# Immunothrombosis shapes the interaction between neutrophils and synovial fibroblasts in rheumatoid arthritis

**DOI:** 10.1101/2025.11.12.25339883

**Authors:** Evangelos Papadimitriou, Anastasia-Maria Natsi, Victoria Tsironidou, Evgenios Eftalitsidis, Maarten van der Linden, Kelsy Waaijenberg, Eric Meldrum, Panagiotis Liakopoulos, Konstantinos Tilkeridis, Athanasios Ververidis, Efthymios Iliopoulos, Maria Koffa, Petros Kolovos, Renato Chirivi, Konstantinos Ritis, Ioannis Mitroulis, Charalampos Papagoras

**Affiliations:** First Department of Internal Medicine, University Hospital of Alexandroupolis, Democritus University of Thrace, Alexandroupolis, Greece; Laboratory of Molecular Hematology, Department of Medicine, Democritus University of Thrace, Alexandroupolis, Greece; Laboratory of Cell Biology, Proteomics and Cell Cycle, Department of Molecular Biology and Genetics, Democritus University of Thrace, Alexandroupolis, Greece; Research and Development, Citryll B.V., Oss, The Netherlands; Department of Molecular Biology and Genetics, Democritus University of Thrace, Alexandroupolis, Greece; Academic Orthopaedics Department, University Hospital of Alexandroupolis

**Author notes:** orresponding author. These authors have contributed equally to this work (Evangelos Papadimitriou, Anastasia-Maria Natsi, Ioannis Mitroulis, Charalampos Papagoras).

**Keywords:** rheumatoid arthritis, fibroblast-like synoviocytes, neutrophil extracellular traps, tissue factor, thrombin

## Abstract

Rheumatoid arthritis (RA) is a chronic autoimmune disease marked by persistent synovial inflammation, yet the processes driving disease progression are not completely understood. Here, we examined the role of fibroblast-like synoviocytes (FLS) and neutrophils in RA pathophysiology, using primary FLS, neutrophils, and synovial fluid (SF) from RA and osteoarthritis (OA) patients, as well as healthy controls. Our findings demonstrate that FLS and neutrophils drive an immunothrombotic state in RA SF by expressing tissue factor (TF), an effect mediated by JAK1/2 signaling. Furthermore, we showed that RA SF stimulates IL-8 (CXCL8) expression in control FLS through PAR-1 signaling, and this response was attenuated by DNase I treatment and CIT-013, a monoclonal antibody targeting anti-citrullinated histones H2A and H4 in neutrophil extracellular traps (NETs), supporting the hypothesis that the effect is mediated by NETs. Notably, FLS derived from RA patients exhibit enhanced CXCL8 expression, and elevated IL-8 levels were detected in RA SF, both contributing to neutrophil recruitment, a process that could be mitigated through blockade with an anti-CXCL-8 neutralizing antibody. These results suggest an amplification loop in which TF expression, thrombin activity, and NET formation converge to activate FLS, sustain IL-8 mediated neutrophil migration, and perpetuate synovial inflammation, revealing how stromal and immune cells interact to propagate RA pathophysiology.

## Introduction

Rheumatoid arthritis (RA) is a chronic autoimmune disease characterized by persistent synovial inflammation, progressive joint damage, and systemic manifestations (1–3). Fibroblast-like synoviocytes (FLS), the key stromal cells of the joint, play an integral role in the pathogenesis of the disease by contributing to cartilage degradation and immune cell recruitment through the production of proinflammatory cytokines, chemokines and matrix-degrading enzymes (4,5). The inflammatory process is sustained by the dysregulated crosstalk between immune and stromal cells within the synovial microenvironment. Among these, neutrophils play pivotal roles in perpetuating inflammation and tissue damage. Neutrophils, which infiltrate the synovium early in disease, are effector cells of acute inflammation and key drivers of chronic tissue injury, through the formation of neutrophil extracellular traps (NETs) (6,7).

In addition to common inflammatory mediators, recent evidence highlights the role of coagulation factors in RA pathophysiology. Thrombin, a serine protease central in the coagulation cascade, is elevated in RA synovial fluid and serves as a marker of local inflammation (8). Beyond its primary role in clot formation, thrombin can activate FLS and immune cells through protease-activated receptor (PAR) signaling, thereby promoting inflammatory cytokine production (9,10). Tissue factor (TF) expression is also upregulated in RA joints, contributing to local thrombin formation and inflammatory signaling (11). The coagulation cascade intersects with other inflammatory pathways, including the JAK/STAT pathway, enhancing immune cell activation and cytokine expression (12).

Interleukin-8 (IL-8/CXCL8) is a potent neutrophil chemoattractant that plays a critical role in the recruitment of neutrophils into the inflamed joint (13,14). Increased levels of IL-8 have been consistently detected in both synovial fluid and serum of RA patients and are closely associated with disease severity and joint damage (15–17). FLS are a source of IL-8 in the synovial tissue, and its production is amplified by inflammatory stimuli and cell–cell interactions (18). Through IL-8 secretion, FLS not only recruit neutrophils, but also sustain an inflammatory feedback loop that promotes NET formation, exacerbating tissue damage and autoantigen exposure (19).

This study aims to investigate the contribution of thrombin–TF signaling to the inflammatory process in RA, focusing on the crosstalk between neutrophils and FLS.

## Results

### Increased tissue factor activity and thrombin generation in RA synovial fluid

RA is characterized by chronic synovial inflammation and an aberrant activation of coagulation pathways within the joint microenvironment. To this direction, we measured thrombin levels in synovial fluid (SF) from patients with active RA and compared them to osteoarthritis (OA) samples by ELISA, and we observed that the concentration of thrombin was significantly elevated in RA SF compared to OA SF (**Figure S1A**). To the same direction, TF enzymatic activity was increased in RA SF compared to OA SF (**Figure S1B**) and significantly correlated with thrombin levels (**Figure S1C**).

### Fibroblast-like synoviocytes constitute a source of tissue factor in RA

To identify the cellular source of TF in RA SF, we initially performed bioinformatic analysis of publicly available single cell RNA-sequencing data (20), to identify cell subsets that express TF. We observed that among other cell populations (**Figure 1A**), FLS expressed *F3*, the gene encoding TF. Additionally, the frequency of *F3* positive FLS was increased in RA compared to OA (**Figure 1B**).

**Figure 1.**
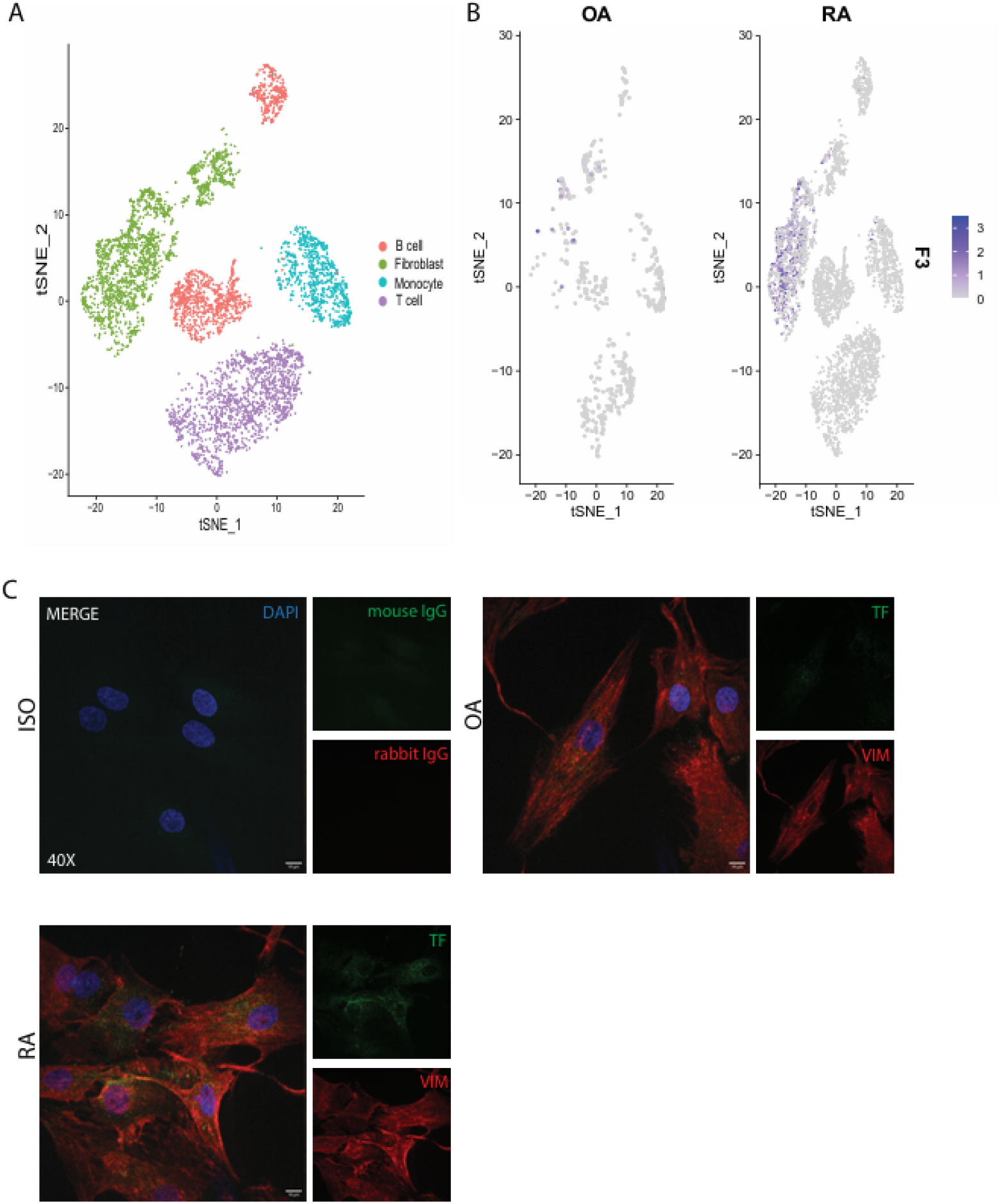
FLS contribute to TF production within the synovial RA environment. **(A)** tSNE plots of scRNAseq data showing synovial cell populations including B cells, fibroblasts, monocytes, and T cells. **(B)** tSNE plots displaying F3 expression in synovial cells derived from OA and RA patients. **(C)** Immunofluorescence staining of primary FLS derived from OA, and RA patients showing TF (green) and Vimentin (red). Corresponding isotype-matched controls were included. One representative example from three independent donors per group are shown. Confocal microscopy; magnification: 40x; scale bar: 10 _μ_m. TF, tissue factor; F3, coagulation factor III (tissue factor gene); VIM, vimentin; FLS, fibroblast-like synoviocytes; ctrl, control subjects; OA, osteoarthritis; RA, rheumatoid arthritis.

To validate these findings, we performed immunofluorescence staining in primary FLS isolated from RA and OA patients, and we observed increased TF expression in FLS from patients with RA (**Figure 1C**).

### Increased neutrophil-derived TF expression correlates with NETosis in RA synovium

Given the abundance of neutrophils in RA SF and their established role in inflammation and immune-mediated thrombosis (21,22), we then focused on neutrophils as a potential source of TF. Immunofluorescence staining **(Figure 2A)** and quantification of *F3* mRNA levels using qPCR studies **(Figure 2B)** revealed increased TF expression in peripheral blood (PB) neutrophils. Similarly, increased levels of TF were observed in synovial fluid neutrophils **(Figure 2C)** from patients with RA. To investigate the extracellular release of TF in the form of NETs, we visualized NET formation using an anti-citrullinated histone H3 (CitH3) antibody. CitH3 was observed within extracellular NET structures of peripheral blood neutrophils (**Figure 3A**) and within the nuclei of SF neutrophils undergoing activation along the NETosis pathway (**Figure 3C**) with some signs of TF colocalization.

**Figure 2.**
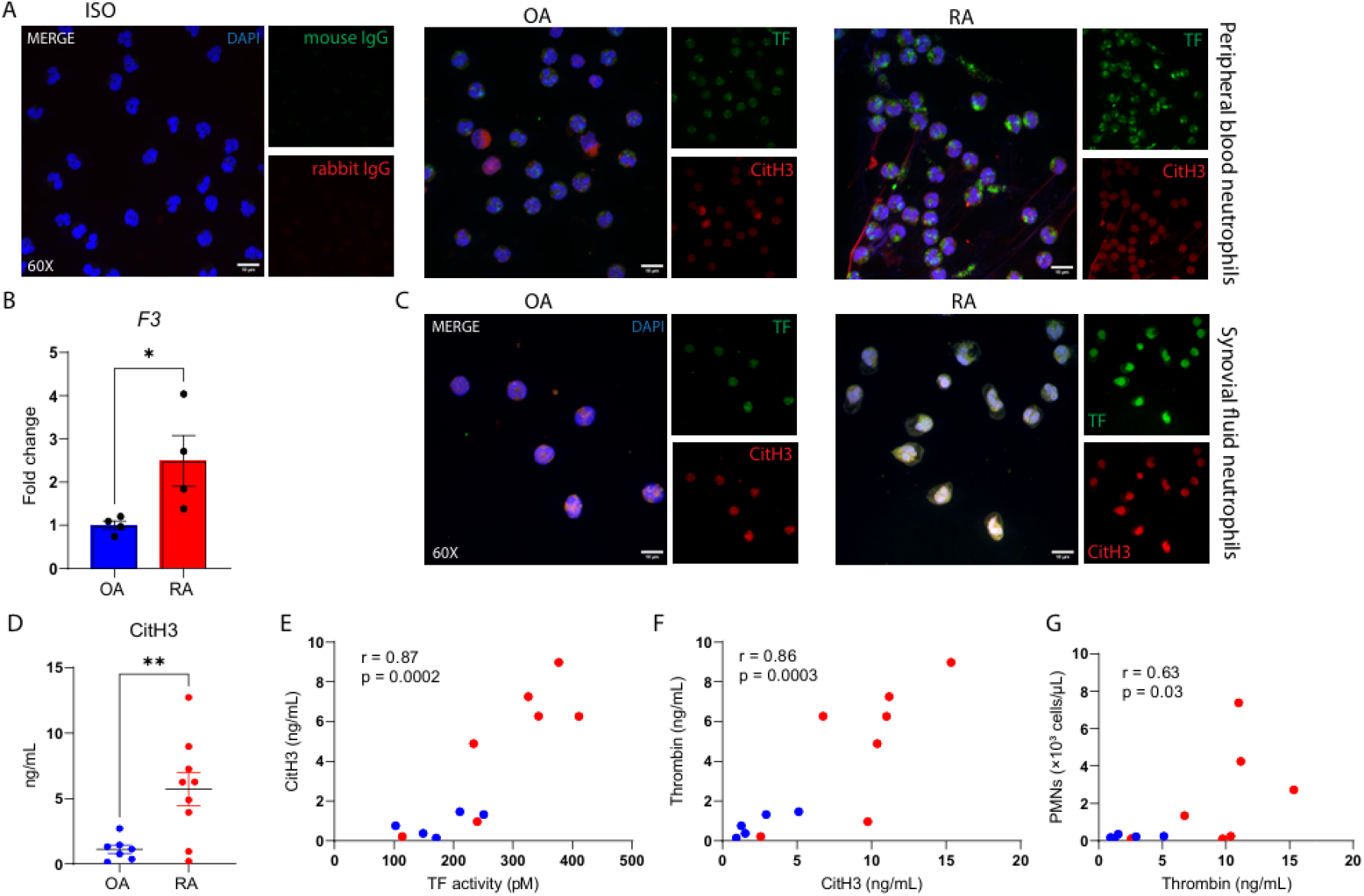
Neutrophil-derived TF and NETs contribute to a proinflammatory environment in RA. **(A)** Immunofluorescence staining in peripheral blood neutrophils from RA and OA patients showing TF (green) and CitH3 (red). Appropriate isotype controls were included. **(B)** Quantification of F3 mRNA by qPCR in peripheral blood neutrophils from OA (n=4) and RA (n=4) patients. Data are expressed as a fold change relative to mean OA transcript levels. (**C)** Immunofluorescence staining of SF neutrophils from OA and RA patients showing TF (green) and CitH3 (red). **(D)** ELISA analysis of CitH3 levels in OA (n=7) and RA (n=9) SF samples. **(E)** Correlation between TF activity and CitH3 levels in OA (n=5) and RA (n=7) SFs. **(F)** Correlation between thrombin levels and CitH3 levels in the same samples. **(G)** Correlation between thrombin concentration and neutrophil counts (×10^3^ cells/μL) in OA and RA SFs. In panels A and B, one representative example from three independent experiments using samples from three different donors is shown. Confocal microscopy; magnification: 60x; scale bar: 10□μm. An unpaired t-test was applied in panels C and D. Pearson correlation was used in panels E-G. Data are presented as mean ± SEM. *p < 0.05, **p < 0.01. TF, tissue factor; CitH3, citrullinated histone H3; RA, rheumatoid arthritis; OA, osteoarthritis; PMNs, polymorphonuclear neutrophils; SF, synovial fluid; NETs, neutrophil extracellular traps.

**Figure 3.**
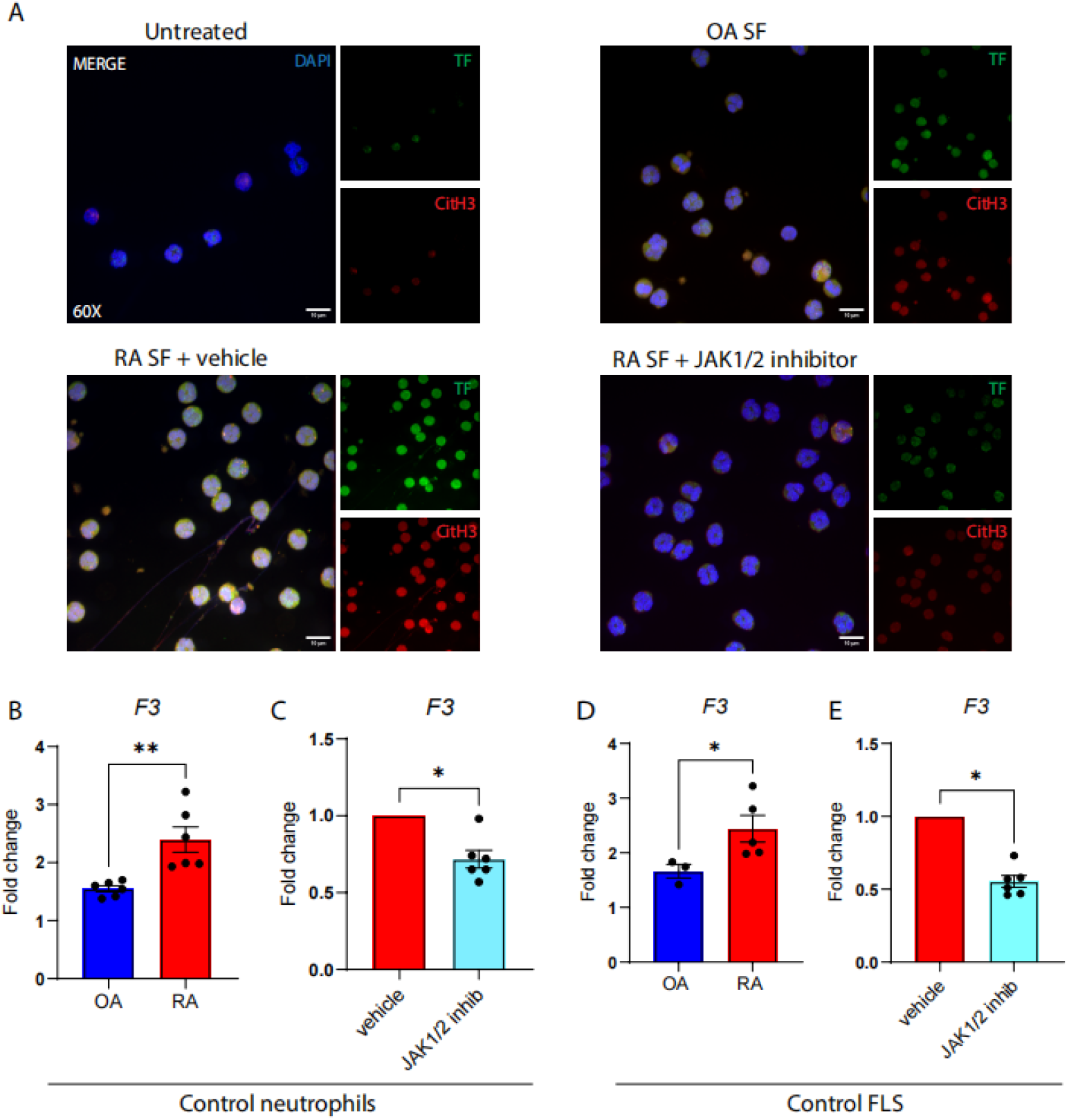
JAK1/2 inhibition attenuates TF expression in RA. **(A)** Immunofluorescence staining of control neutrophils treated with 10% OA or RA SF, with or without pre-treatment with baricitinib. Appropriate vehicle control was used. TF (green) and CitH3 (red) were visualized by confocal microscopy. **(B)** qPCR analysis of F3 mRNA levels in control neutrophils treated with 10% OA or RA SF. Data are expressed as fold change compared with the untreated control group. **(C)** F3 mRNA expression in control neutrophils pre-treated with baricitinib or vehicle control prior to RA SF stimulation. **(D)** F3 mRNA expression in control FLS stimulated with SF from OA (n=3) or RA (n=5) patients. Data are expressed relative to untreated controls. **(E)** F3 mRNA expression in control FLS pre-treated with baricitinib or vehicle control prior to stimulation with RA SF. In panel A, one representative example from three independent experiments is shown. Magnification: 60x; scale bar: 10 μm. Data in panels (B) and (D) were analyzed using an unpaired t-test, and data in panels (C) and (E) using a paired t-test. Data are presented as mean ± SEM. *p < 0.05, **p < 0.01. TF, tissue factor; F3, coagulation factor III (tissue factor gene); CitH3, citrullinated histone H3; SF, synovial fluid; RA, rheumatoid arthritis; OA, osteoarthritis.

Quantification of CitH3 in SF using ELISA revealed significantly higher NET levels in RA SF compared to OA SF (**Figure 2D**). Interestingly, levels of CitH3 were significantly correlated with TF activation and thrombin generation in SF (**Figure 2E, F**). Furthermore, a positive correlation was identified between synovial neutrophil numbers and thrombin concentration (**Figure 2G**), proposing that neutrophil activation and NET formation may contribute to the amplification of the TF – dependent thrombin production within the RA synovial compartment.

### JAK1/2 inhibition attenuates tissue factor expression in RA

Several cytokines are implicated in RA pathogenesis that signal through the JAK/STAT pathway, while JAK inhibition is an efficacious treatment of RA (23). For this reason, we studied whether blocking JAK/STAT signaling attenuates the enhanced expression of TF in RA. To this end, control neutrophils were stimulated with SF from OA and RA patients in the presence or absence of the JAK1/2 inhibitor baricitinib. JAK1/2 inhibition significantly reduced TF expression in neutrophils exposed to RA SF, as demonstrated by decreased TF protein levels (**Figure 3A**) and downregulation of *F3* expression (**Figure 3B, C**). Similarly, JAK1/2 inhibition downregulated the *F3* expression in control FLS treated with SF from patients with RA (**Figure 3D, E**).

### IL-8 from FLS promotes neutrophil recruitment in RA SF

The significant correlation between the activation of the coagulation cascade and synovial fluid neutrophil numbers prompted us to further focus on CXCL8 (IL-8), a key neutrophil chemoattractant and activator (24). IL-8 levels were significantly elevated in the SF from patients with RA compared to OA, as assessed by ELISA (**Figure 4A**). To further assess the possible role of IL-8 in neutrophil infiltration, we further compared IL-8 levels in the SF from polymorphonuclear (PMN)-rich and PMN-poor RA SF samples and observed significantly higher IL-8 levels in PMN-rich samples (**Figure 4B**). Moreover, neutrophil counts showed a positive correlation with IL-8 concentrations (**Figure 4C**). Additionally, IL-8 levels were significantly correlated with the levels of CitH3 (**Figure 4D**), supporting a link between neutrophil recruitment and activation.

**Figure 4.**
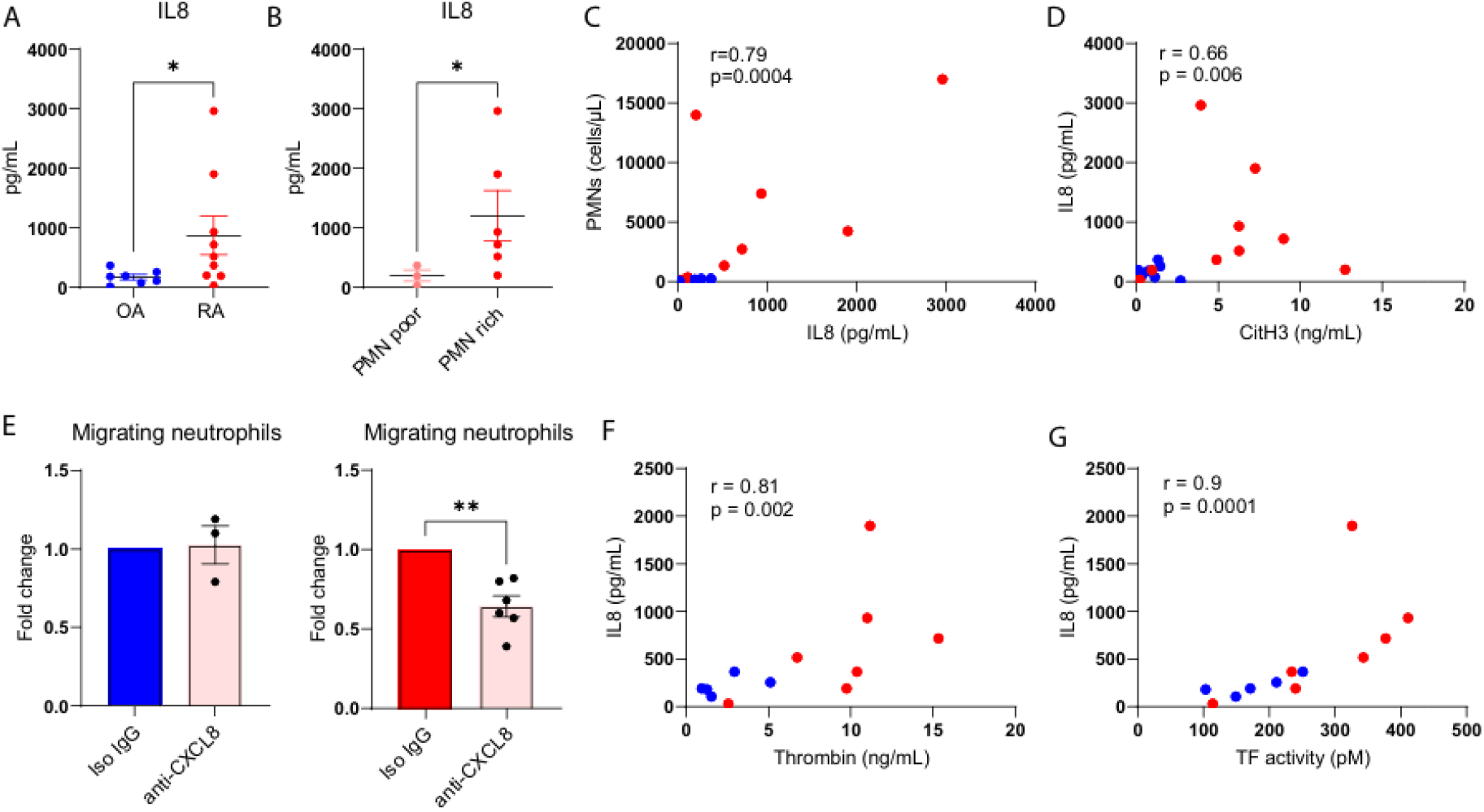
IL-8 links neutrophil activation, NET formation, and TF activity in the inflamed RA synovium. **(A)** ELISA quantification of IL-8 levels in SF samples from OA (n=7) and RA (n=9) patients. **(B)** Comparison of IL-8 ELISA levels in RA SF samples with high versus low PMN counts. **(C)** Correlation between IL-8 ELISA levels and neutrophil counts (cells/μL) in OA and RA SFs. **(D)** Correlation between IL-8 and CitH3 ELISA levels in the same SF samples. **(E)** Transwell chemotaxis assays assessing neutrophil migration toward OA (n=3) and RA (n=5) SF, with or without pre-treatment using a neutralizing anti-CXCL8 antibody or IgG control. **(F)** Correlation between IL-8 and thrombin concentrations in SF samples. **(G)** Correlation between IL-8 levels and TF enzymatic activity in the same samples. Panels A and B were analyzed using the Mann–Whitney U test; panel E was analyzed using a paired t-test. Spearman correlation was used for panels C, D, F, and G. Data are presented as mean ± SEM. *p < 0.05, **p < 0.01. IL-8, interleukin-8; Chemokine (C-X-C Motif) Ligand 8, CXCL8; CitH3, citrullinated histone H3; TF, tissue factor; RA, rheumatoid arthritis; OA, osteoarthritis; IgG, isotype control antibody; PMN, polymorphonuclear neutrophil; SF, synovial fluid. Based on these findings, we next assessed the IL-8 expression in primary FLS. RA FLS showed significantly higher IL-8 expression at both protein and mRNA levels compared to OA FLS (**Figure 5A, B**), indicating that FLS are a potential source of IL-8 in the RA joint. To further study the regulation of IL-8 in FLS, we stimulated control FLS with SF from RA and OA patients. RA SF induced a markedly higher increase in CXCL8 expression compared to OA SF (**Figure 5C**), suggesting that inflammatory mediators present in RA SF enhance IL-8 production by FLS. Next, we performed a transwell neutrophil chemotaxis assay using conditioned media from RA FLS cultures. Preincubation of RA FLS supernatants with a CXCL8-neutralizing antibody significantly reduced neutrophil migration (**Figure 5D**).

**Figure 5.**
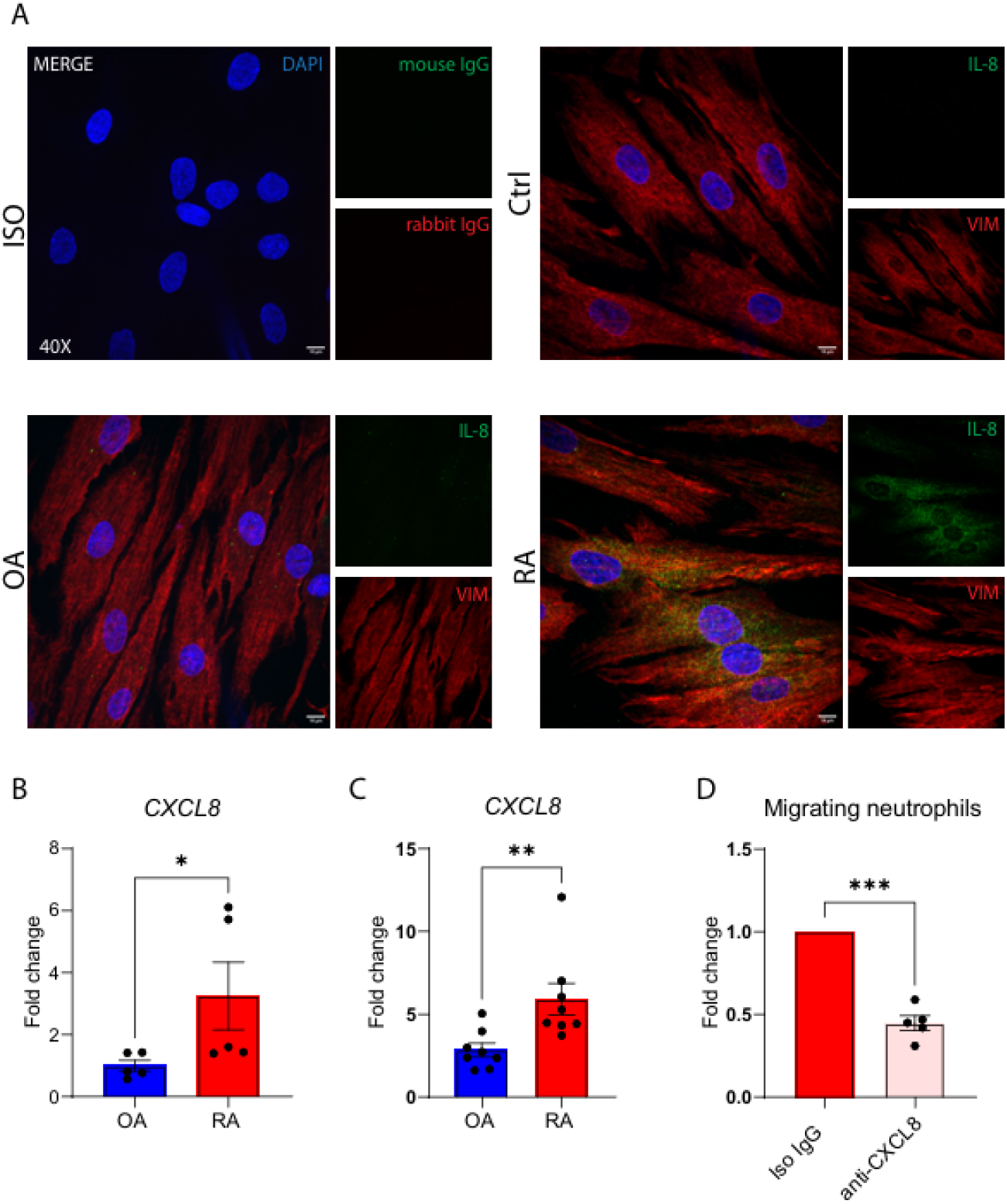
FLS-derived IL-8 drives neutrophil recruitment in the RA joint. **(A)** Immunofluorescence staining of primary FLS from control subjects, OA, and RA patients showing IL-8 (green) and vimentin (red). Appropriate isotype controls were used. **(B)** RT-qPCR analysis of CXCL8 mRNA expression in FLS from OA (n=5) and RA (n=5) patients. Data expressed as fold change relative to the mean of OA samples. **(C)** CXCL8 mRNA levels in control FLS treated with 10% OA (n=8) or RA (n=8) SF. Data are expressed as fold change relative to untreated controls. **(D)** Transwell chemotaxis assay showing neutrophil migration in response to supernatants from RA FLS (n=5), with or without treatment using a neutralizing anti-CXCL8 antibody or IgG control. Statistical analysis was performed using Mann– Whitney U tests for panels B and C, and a paired t-test for panel D. Data are presented as mean ± SEM. *p < 0.05; **p < 0.01; ***p < 0.001. IL-8, interleukin-8; CXCL8, chemokine (C-X-C motif) ligand 8; VIM, vimentin; FLS, fibroblast-like synoviocytes; ctrl, control subject; OA, osteoarthritis; RA, rheumatoid arthritis; IgG, isotype control antibody; SF, synovial fluid.

To functionally assess the chemoattracting potential of IL-8, we conducted a transwell migration assay using control neutrophils. Blocking of IL-8, using a CXCL8-neutralizing antibody, had no significant effect on neutrophil migration compared to isotype control-treated SF when OA SF was used as a chemoattractant (**Figure 4E**). In contrast, preincubation of RA SF with a CXCL8-neutralizing antibody markedly reduced neutrophil migration relative to isotype control-treated SF (**Figure 4E**). These findings confirm that IL-8 is an important mediator of neutrophil recruitment in RA. Furthermore, IL-8 concentrations were positively correlated with thrombin levels in the SF (**Figure 4F**), and with TF activity in SF (**Figure 4G**).

### NET components and thrombin/PAR-1 signaling amplify *CXCL8* expression in FLS

To identify mediators that promote IL-8 production by FLS in RA, we stimulated control FLS with RA SF. RA SF stimulation led to a marked increase in *CXCL8* mRNA expression in control FLS, which was significantly attenuated by DNase I treatment, implicating extracellular DNA as a key inducer of IL-8 (**Figure 6A**). Consistent with this, ELISA measurements revealed significantly higher levels of citrullinated histones H2A and H4 (CitH2A/H4) in RA compared to OA SF (**Figure 6B**), indicating an increased presence of NET-derived components in RA SF. Based on these observations, we next investigated whether these citrullinated histones could modulate fibroblast activation. To this end, we treated RA SF with the anti-citrullinated histone antibody CIT-013 (25), which selectively binds to citrullinated histone H2a and citrullinated histone H4, before FLS stimulation. Neutralization by CIT-013 significantly reduced *CXCL8* mRNA levels compared to isotype IgG1 controls (**Figure 6C**), suggesting that citrullinated histones contribute to the stimuli associated with RA SF-induced *CXCL8* expression in FLS. The same RA and OA SF samples used in the ELISA in Figure 6B were also applied for the FLS stimulation in Figure 6C, linking the elevated CitH2A and CitH4 content in RA SF with the *CXCL8* expression observed in FLS.

**Figure 6.**
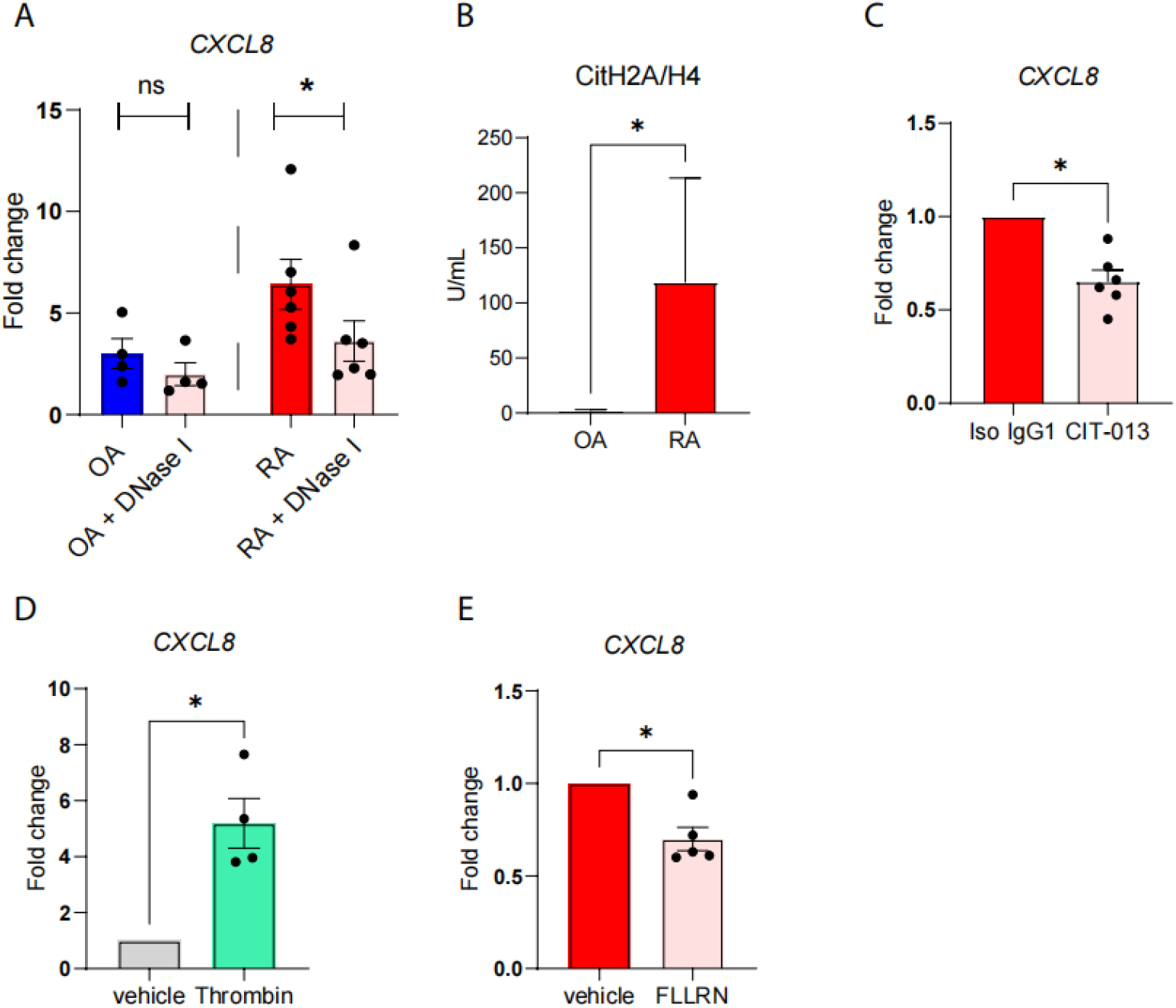
NETs and thrombin/PAR-1 signaling drive CXCL8 expression by FLS in RA synovium. **(A)** CXCL8 mRNA expression in control FLS stimulated with 10% OA or RA SF, with or without DNase I treatment. Data are presented as fold change relative to untreated FLS. **(B)** Citrullinated H2A/H4 levels in OA (n=4) and RA (n=6) SFs.**(c)** CXCL8 mRNA levels in control FLS stimulated with 10% RA SF pre-treated with the CIT-013 or IgG1 control. **(D)** CXCL8 mRNA expression in control FLS following thrombin stimulation compared to vehicle control. **(E)** CXCL8 mRNA expression in control FLS stimulated with 10% RA SF in the presence or absence of the PAR-1 antagonist FLLRN. Wilcoxon matched-pairs signed-rank tests were used for panels A and C; Mann–Whitney U test for panels B and E; and paired parametric t-test for panel D. Data are presented as mean ± SEM. *p < 0.05. CXCL8, chemokine (C-X-C motif) ligand 8; FLS, fibroblast-like synoviocytes; RA, rheumatoid arthritis; OA, osteoarthritis; SF, synovial fluid; NETs, neutrophil extracellular traps; IgG1, isotype control antibody; CIT-013, monoclonal antibody against citrullinated histones; CitH2A/H4, citrullinated histones H2A and H4.

We next evaluated the contribution of thrombin/PAR-1 signaling to this response, given that thrombin levels were positively correlated with IL-8 concentrations in RA SF. Stimulation with purified thrombin alone was sufficient to induce *CXCL8* mRNA expression in control FLS (**Figure 6D**). Additionally, inhibition of PAR-1 with the specific antagonist peptide FLLRN significantly reduced *CXCL8* mRNA expression in RA SF–stimulated FLS compared with the vehicle control (**Figure 6E**).

## Discussion

This study provides evidence for a bidirectional link between the coagulation system and neutrophil activation in RA. We demonstrate that FLS and neutrophils express TF and contribute to the activation of the coagulation cascade in RA synovial fluid. We further provide evidence supporting that the generated thrombin, as well as NET structures, induce the production of the neutrophil chemotactic factor IL-8 production in FLS which might trigger neutrophil influx into the inflamed joint. A key finding of our study is that TF activity and thrombin levels are markedly elevated in RA SF, corroborating previous reports of aberrant coagulation activation in the RA joint environment (25,26). We further identify neutrophils and FLS as important cellular sources of TF.

Interleukin-1β and TNF-α are well-characterized drivers of FLS activation, promoting the expression of IL-6, IL-8 and MMPs, which sustain local synovial inflammation and joint damage (27). However, the direct role of FLS in fueling the coagulation cascade in RA has not been clearly demonstrated. On the other hand, the risk of cardiovascular and thromboembolic events is increased in RA patients, particularly those with high disease activity (28,29). Thus, our results suggest that activated FLS are an additional cellular contributor to the pro-coagulant state directly proportional to the state of inflammation. Moreover, the role of NETs in promoting coagulation is well established. Previous work from our group has demonstrated that NETs in ANCA-associated vasculitis contribute to thromboinflammation via TF exposure and the release of TF-bearing microparticles (30). The detection of CitH3-positive neutrophils expressing TF within the joint, alongside the positive correlations between CitH3, thrombin, and TF activity levels, suggest a mechanistic link between NETosis and neutrophil-mediated TF expression in RA, as well.

Our data further provide evidence that the inflammatory milieu of RA SF is sufficient to induce TF expression in FLS *via* a JAK1/2-dependent mechanism. These findings expand the current understanding of the contribution of FLS in joint inflammation by highlighting their role in local hypercoagulability and reinforce the view of FLS as dynamic stromal cells that are not only effectors of the inflammatory process, but also actively shape the inflammatory environment of the RA joint (31). Notably, our data also show that JAK1/2 inhibition significantly reduces TF expression in neutrophils, as well, indicating that cytokine-driven signaling may serve as a critical convergence point between inflammatory activation, NET formation, and coagulation in RA.

Elevated IL-8 levels in RA SF were previously shown to correlate with disease activity and neutrophil infiltration (17,24,32). Our findings provide a mechanism for this correlation by demonstrating that FLS are a major source of IL-8, and that RA SF and NET-derived signals potently stimulate IL-8 production. Particularly, the RA SF enriched in citrullinated histones, key components of NETs, was capable of directly inducing *CXCL8* mRNA expression in FLS, an effect reversed by the NET targeting antibody CIT-013. These findings support the idea that NET-derived citrullinated histones present in RA SF not only contribute to tissue damage but also act as paracrine activators of stromal cells, thereby perpetuating synovial inflammation (19,33).

In addition to NETs, we identify thrombin/PAR-1 signaling as a crucial inducer of IL-8 expression in FLS. Thrombin has previously been shown to activate FLS *via* PAR-1 and induce IL-6 production, promoting pro-inflammatory cytokine secretion (34). Additionally, thrombin has been previously reported to stimulate IL-8 expression in dermal fibroblasts through PAR-1–dependent mechanisms (35). Our findings confirm and extend these observations by demonstrating that both purified thrombin and RA SF–mediated PAR-1 activation significantly enhance IL-8 expression in FLS. Importantly, PAR-1 blockade reduced IL-8 expression, confirming a direct contribution of thrombin to neutrophil recruitment signals. This suggests that thrombin sustains synovial inflammation by promoting FLS activation and subsequent neutrophil recruitment.

In conclusion, our findings support a model in which FLS and neutrophils cooperate to amplify synovial inflammation through a feedback loop involving NET formation, TF expression, thrombin generation and IL-8 production. In this model, RA SF acts as a potent inflammatory amplifier by inducing TF expression in both FLS and neutrophils through JAK/STAT pathway mediated IL-8 expression in FLS via PAR-1 signaling. In turn, NETs, enriched in citrullinated proteins, further activate FLS to produce IL-8 and promote neutrophil recruitment.

## Data Availability

All data produced in the present study are available upon reasonable request to the authors

## Authors’ Contributions

EP: Investigation, Methodology, Visualization, Writing – original draft, Writing – review & editing. A-MN: Investigation, Methodology, Visualization, Writing – original draft, Writing – review & editing. VT: Investigation, Methodology. EE: Visualization. ML: Methodology, Writing – review & editing. KW: Methodology, Writing – review & editing. EM: Review. PL: Methodology, Visualization. KT: Methodology. AV: Methodology. GD: Methodology. MK: Writing – review & editing. PK: Methodology, Writing – review & editing. RC: Investigation, Methodology, Review & editing. KR: Conceptualization, Writing – review & editing. IM: Conceptualization, Funding acquisition, Project administration, Supervision, Writing – original draft, Writing – review & editing. CP: Conceptualization, Funding acquisition, Project administration, Supervision, Writing – original draft, Writing – review & editing.

