## Supplementary methods, Table and Figures for "Immunothrombosis shapes the interaction between neutrophils and synovial fibroblasts in rheumatoid arthritis"

**Supplementary Materials and Methods**

**1. Patients and sampling**

All participants were adults under routine clinical care and provided written informed consent. In total 9 RA patients, 8 OA patients and 5 control subjects were recruited. From enrolled patients, peripheral blood and/ or SF were collected. Synovial tissue was additionally obtained from OA and RA patients during elective total knee arthroplasty. Control subjects consisted of individuals undergoing arthroscopy for recent mechanical trauma but no active joint inflammation. The study protocol was approved by the Ethics Committee of University Hospital of Alexandroupolis. The baseline clinical characteristics of patients and samples used in experimental procedures are summarized in **Supplementary Table 1**.

**2.** **Collection of synovial fluid (SF) and isolation of peripheral blood- and SF-derived neutrophils**

SF samples were obtained from patients undergoing therapeutic joint aspiration by an experienced rheumatologist under aseptic conditions. Immediately after collection, samples were transferred into sterile EDTA or sodium citrate vacutainer tubes. SF was centrifuged at 800 × g for 10 min at 4 °C to remove cells and debris. The resulting supernatant was either used immediately for experiments or aliquoted and stored at –80 °C until further analysis.

Peripheral blood was obtained in heparin-coated tubes, and circulating neutrophils were isolated using a double-density Histopaque gradient centrifugation method (densities: 1.119 g/mL and 1.077 g/mL; Sigma-Aldrich, St. Louis, MO, USA). Samples were centrifuged at 700×g for 35 minutes at room temperature (20–25°C). Following isolation, neutrophils were washed once with phosphate-buffered saline (1× PBS, Biosera) and pelleted by centrifugation at 200×g for 12 minutes. Cells were then resuspended at the required concentration. Neutrophil purity was routinely ≥ 98%.

SF was collected from patients with RA or OA and diluted 1:1 with 1× PBS (Biosera). Diluted SF was layered onto a double-density Histopaque gradient (densities: 1.119 g/mL and 1.077 g/mL; Sigma-Aldrich) and centrifuged at 700×g for 35 minutes at 20–25°C without brake. Neutrophils were collected from the interface between the two gradients, washed once with 1× PBS, and centrifuged at 200×g for 12 minutes. Cells were resuspended at the desired concentration and used within two hours of isolation.

**3. Isolation, culture, and characterization of human primary fibroblast-like synoviocytes (FLS)**

Synovial tissue was obtained from five control subjects undergoing orthopedic procedures unrelated to inflammatory joint disease, while SF was collected from five patients with RA and five with OA during diagnostic arthrocentesis. All procedures were performed in accordance with ethical guidelines and informed consent.

Fresh synovial tissue was rinsed with sterile PBS (Biosera), minced (~1 mm³), and digested with 0.1% trypsin (Thermo Fisher Scientific) for 30 min at 37 °C followed by 0.1% type I collagenase (Sigma-Aldrich) in DMEM (Biosera) supplemented with 10% FBS for 2 h at 37 °C under constant agitation. Digested fragments were washed, seeded into flasks, and cultured in DMEM supplemented with 10% FBS (Gibco), 100 U/mL antibiotic/antimycotic, and 1% non-essential amino acids (Thermo Fisher Scientific).

Synovial fluid samples were processed within 1–2 h, diluted 1:1 with PBS, and centrifuged (400×g, 10 min). Cell pellets were resuspended in complete DMEM, seeded, and incubated for 48 h at 37 °C, 5% CO₂, after which non-adherent cells were removed. Adherent FLS were expanded with medium changes every 3–4 days.

For both tissue- and fluid-derived FLS, adherent cells were expanded and passaged at 80–90% confluence, and passages 4–8 were used in experiments to ensure phenotypic stability (1,2).

FLS identity was confirmed based on their spindle-shaped morphology and characterized by immunofluorescence staining using established mesenchymal markers. Cells were stained for CD90 (THY1), desmin, and vimentin to confirm their fibroblastic phenotype. FLS were consistently positive for CD90 and vimentin, while desmin expression was absent, consistent with their non-muscle, mesenchymal identity.

**4. Stimulation and inhibition studies in cultured cells**

Peripheral blood neutrophils (1.5 × 10⁶/well) and primary FLS (1.5 × 10⁵/well) were seeded in 6-well plates (Corning Incorporated) and cultured in complete RPMI or DMEM, respectively.

Neutrophils were stimulated with 10% SF from OA or RA patients at 37 °C, 5% CO₂ for 90 min (mRNA) or 3.5 h (protein). To examine JAK signaling, cells were pre-treated with the JAK1/2 inhibitor Baricitinib (2.5 nM; 16707; Cayman Chemical) for 30 min before SF stimulation.

FLS were stimulated with 10% SF (OA or RA) or thrombin (1 U/mL; 605195; Sigma-Aldrich) for 3 h. To investigate signaling pathways, cells were pre-treated with the JAK1/2 inhibitor Baricitinib (2.5 nM; 16707; Cayman Chemical) or the PAR1 antagonist FLLRN (300 μM; AS-60678; Anaspec). Appropriate vehicle controls were included. To assess the contribution of NETs, SF samples were pre-treated with DNase I (10 U/mL; EN0525; Thermo Fisher) or with the anti-citrullinated histone H2A/H4 antibody CIT-013 (10 μg/mL; Citryll) or isotype IgG1 control (10 μg/mL; C0001-5; CrownVivo).

**5. RNA isolation, cDNA synthesis and RT-qPCR**

Total RNA was extracted from primary FLS and peripheral blood neutrophils using TRIzol reagent (15596026; Thermo Fisher Scientific) according to the manufacturer’s instructions. Equal amounts of RNA were used for complementary DNA (cDNA) synthesis, which was carried out using the PrimeScript™ RT Reagent Kit (RR037A; Takara) following the provided protocol. Quantitative real-time PCR (RT-qPCR) was performed to assess the expression of *C-X-C Motif Chemokine Ligand 8/IL8* (*CXCL8*) and *Coagulation Factor III/Tissue Factor* (*F3*) using KAPA SYBR FAST qPCR Master Mix (2X) (KK4602; KAPA Biosystems). Glyceraldehyde 3-phosphate dehydrogenase (GAPDH) served as internal control for normalization. Relative gene expression was calculated using the 2^–ΔΔCt^ method.

**6. Immunofluorescence staining of neutrophils and FLS**

Freshly isolated neutrophils (1.5 × 10⁵) from peripheral blood of RA, OA, and control subjects, as well as from RA and OA synovial fluid, were seeded on poly-D-lysine–coated coverslips and incubated for 3.5 h in RPMI-1640 (Biosera) with 2% FBS. Cells were fixed with 4% paraformaldehyde (15 min, RT), permeabilized with 0.5% Triton X-100 (1 min), and blocked with 6% normal goat serum (31872; Invitrogen). Primary antibodies included anti-TF (1:150; 4509; American Diagnostica) and anti-CitH3 (1:1000; ab281584; Abcam); secondary antibodies were CF488A goat anti-mouse (20956; Biotium) and CF594 goat anti-rabbit (20955; Biotium). Nuclei and extracellular DNA were counterstained with DAPI (D9542; Sigma-Aldrich). Isotype controls were included in all experiments.

Primary FLS were seeded into 8-well chamber slides (Ibidi) and stained with antibodies against IL-8 (15 μg/mL; MAB208-100; R&D Systems), desmin (1:200; ab15200; Abcam) and vimentin (1:250; ab92547; Abcam), and TF (1:150 dilution; 4509; American Diagnostica). The same secondary antibodies were used. Nuclei were counterstained using a DAPI-containing mounting medium (50011; Ibidi) (3,4).

Imaging was performed on an Andor Revolution spinning-disc confocal system (CSU-X1 scan head on Olympus IX81), equipped with an Andor Zyla 4.2 sCMOS camera and controlled by Andor iQ software (v3.6.5). Images were acquired with 40× (0.95 NA) and 60× (1.42 NA) air objectives at the Bioimaging-DUTH facility.

**7. Tissue factor (TF) activity assay**

TF activity in SF samples from OA and RA patients was quantified using the Tissue Factor Human Chromogenic Activity Assay Kit (ab108906; Abcam), following the manufacturer’s instructions (5). The assay is based on the ability of the TF/FVIIa complex to convert factor X to activated factor Xa. The resulting enzymatic activity is measured by a chromogenic substrate, with the increase in absorbance being directly proportional to TF activity.

**8. Thrombin ELISA**

Thrombin levels in RA and OA SF samples were quantified using the Human Thrombin AssayMax ELISA Kit (ET4010-1; Assaypro) according to the manufacturer’s instructions. SF samples were thawed on ice, diluted in assay buffer, and analyzed in pre-coated 96-well plates. Absorbance was measured at 450 nm (Byonoy).

**9. Citrullinated Histone H3 (CitH3) ELISA**

CitH3 levels were measured in SF samples from RA and OA patients using a CitH3 ELISA kit (501620; Cayman Chemical), following the manufacturer’s instructions. Samples were appropriately diluted, added to pre-coated ELISA plates, and processed according to the kit protocol. Absorbance was read at 450 nm using a microplate reader, and concentrations were calculated from the standard curve.

**10. Citrullinated Nucleosome ELISA and hyaluronidase treatment of SF**

To reduce the viscosity of SF samples prior to ELISA analysis, a hyaluronidase pretreatment was performed. SF samples were first centrifuged at room temperature (RT) at 16,900 × g to remove residual cells. Supernatants were diluted 1:10 in Dulbecco’s phosphate-buffered saline (DPBS) and treated with hyaluronidase (H3506; Sigma-Aldrich) at a final concentration of 100 U/mL. Samples were vortexed and incubated for 1 h at 37 °C with gentle agitation (500 rpm). The pre-treated SF samples were subsequently used for the quantification of citrullinated nucleosomes by ELISA.

For the Citrullinated Nucleosome ELISA, NUNC Maxisorp flat-bottom 96-well plates were coated overnight at 4 °C with 850 ng/mL polyclonal rabbit anti-histone H3 (citrulline R2 + R8 + R17) antibody (ab281584; Abcam) in 1× PBS. Wells were washed three times with PBS containing 0.05% (v/v) Tween™ 20 (PBS-Tween) and blocked with PBS-Tween containing 1% (w/v) bovine serum albumin (PBS-Tween-BSA) for 2 h at RT. After removal of the blocking solution, hyaluronidase-pretreated SF samples were added and incubated for 1 h at RT with gentle agitation (450 rpm). Wells were then washed three times with PBS-Tween and incubated with 400 ng/mL monoclonal mouse anti-citrullinated histones H2A and H4 (mouse version of CIT-013) diluted in PBS-Tween-BSA for 1 h at RT with gentle agitation. After three washes, wells were incubated with 76.9 ng/mL HRP-conjugated polyclonal goat anti-mouse immunoglobulin antibody (Dako, P0447) in PBS-Tween-BSA for 1 h at RT with gentle agitation. Following three final washes, 3,3′,5,5′-tetramethylbenzidine (TMB; 74931569A; Thermo Fisher Scientific,) substrate was added and incubated for 10 min at RT in the dark. The reaction was stopped with an equal volume of 2 M H₂SO₄ (1007311000; Merck), and absorbance was measured at 450 nm (reference 620 nm) using a Tecan Infinite F50 plate reader controlled by Magellan F50 software (version 7.2).

**11. Interleukin-8** (**IL-8) ELISA**

IL-8 levels were quantified in cell-free SF and FLS culture supernatants from RA and OA patients using a human IL-8 ELISA kit (3560-1HP-1; MABTECH), according to the manufacturer’s instructions. SF was precleared by centrifugation to remove cells and debris, aliquoted, and stored at −80 °C until analysis. For analysis related to neutrophil content, SF samples were categorized as polymorphonuclear (PMN)-rich (mean cell count: 7785 cells/μL, range: 1345-14.000) or PMN-poor (mean cell count: 150 cells/μL, range: 100-240) based on differential neutrophil counts obtained at collection.

**12. *In vitro* transwell migration assay (chemotaxis assay)**

Neutrophil chemotaxis was assessed using a fluorimetric 96-well QCM™ Chemotaxis Cell Migration Assay with 3 μm pore inserts (ECM515; Merck Millipore), following the manufacturer’s protocol. The lower chambers were filled with low-serum complete DMEM (serving as a negative control), supernatants derived from OA or RA FLS, or SF from OA and RA patients diluted to 10% in DMEM. Freshly isolated human peripheral blood neutrophils (0.2–2 × 10⁶ cells/mL) were suspended in low-serum complete DMEM and added to the upper chambers.

After incubation for 2.5 hours at 37°C in a 5% CO₂ atmosphere, neutrophil migration was quantified by measuring fluorescence using a PerkinElmer Enspire plate reader (Waltham, MA, USA) with excitation/emission wavelengths set at 480/520 nm. Migration toward FLS supernatant was assessed in five independent experiments, and migration toward SF was assessed in six independent experiments using neutrophils from different healthy donors. The fluorescence intensity of migrated cells was calculated for each condition.

To assess the role of IL-8/CXCL8 in neutrophil chemotaxis, a neutralizing mouse monoclonal anti-IL-8 antibody (2 μg/mL; MAB208-100, R&D Systems) was added to selected conditions. A mouse polyclonal IgG antibody (2 μg/mL; ab37355, Abcam) was used as an isotype control in neutralization assays.

**13. Single cell RNA seq**

Publicly available filtered single-cell RNA sequencing (scRNA-Seq) data from Zhang et al. were processed using Seurat (v4.3.0.1) for normalization, scaling, and visualization (6,7).

**14. Statistical analysis**

Statistical analysis was performed with GraphPad Prism software (version 9.0, San Diego, CA, USA). Data are expressed as the mean ± standard error of the mean (SEM). Two-group comparisons were analyzed with an unpaired, two-tailed Student’s t-test when data were normally distributed; otherwise, a two-tailed Mann–Whitney U test was used. For paired data, we applied a paired, two-tailed Student’s t-test when the distribution of paired differences was normal, or the Wilcoxon matched-pairs signed-rank test when it was not. Linear associations were evaluated using Pearson’s correlation coefficient (r), or Spearman’s rank correlation for non-normal or non-linear relationships. Statistical significance was defined as *p<0.05, **p<0.01, ***p<0.001, ****p<0.0001.

**Supplementary figures**

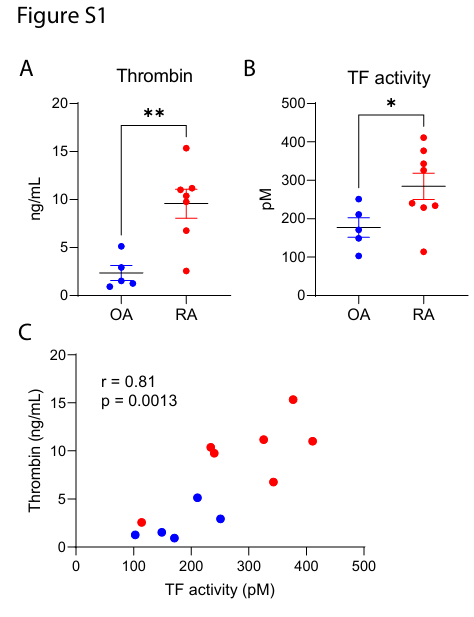

**Figure S1**. Thrombin and TF enzymatic activity are elevated in RA SF. (A) Thrombin levels measured by ELISA in OA (n=5) and RA (n=7) SF samples. (B) TF activity determined by chromogenic assay in OA (n=5) and RA (n=8) SF samples. (C) Correlation analysis between thrombin concentration and TF activity in OA/RA SFs. An unpaired parametric t-test was applied in panels A and B. Pearson correlation analysis was used in panel C. Data are expressed as mean ± SEM. **p < 0.01, *p < 0.05. TF, tissue factor; RA, rheumatoid arthritis; OA, osteoarthritis; SF, synovial fluid.

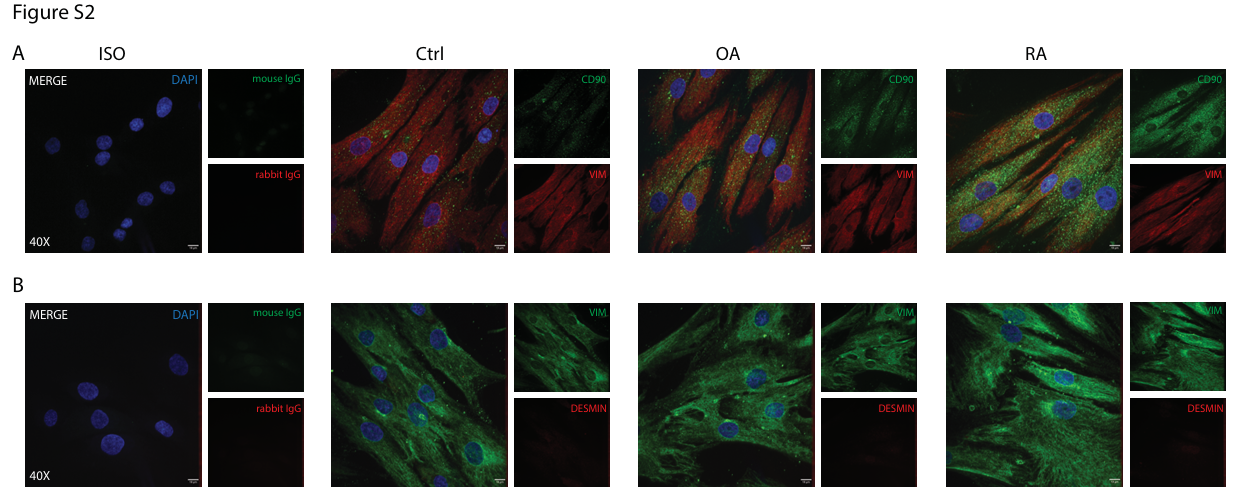

**Figure S2**. Phenotypic characterization of human primary FLS derived from controls, OA, and RA patients. **(A)** Immunofluorescence staining for CD90 (Thy-1, green) and Vimentin (red) in FLS from control subjects, OA, and RA patients. **(B)** Co-staining for Desmin (red) and Vimentin (green). Representative images from five independent donors per group are shown. Confocal microscopy; magnification: 40x; scale bar: 10 μm. CD90, cluster of differentiation 90; VIM, vimentin; ctrl, control subject; OA, osteoarthritis; RA, rheumatoid arthritis; FLS, fibroblast-like synoviocytes.

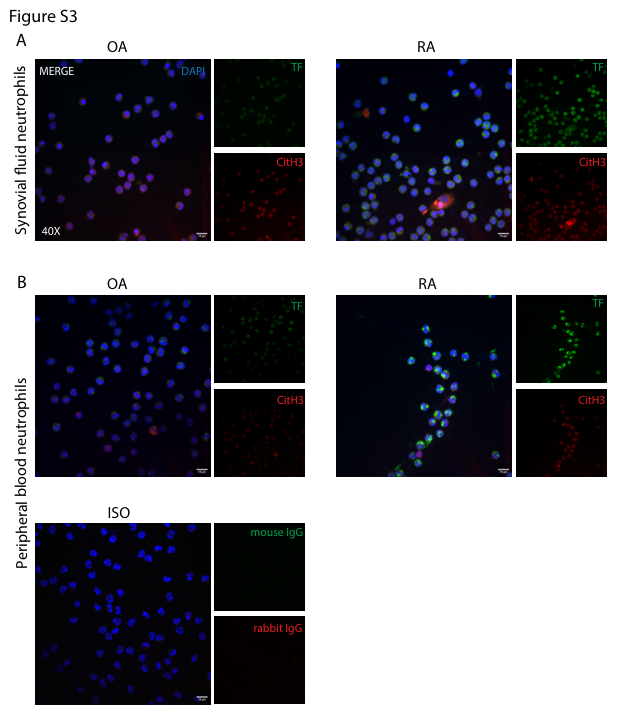

**Figure S3**. Immunofluorescence analysis of TF and CitH3 in SF and peripheral blood neutrophils from OA and RA patients. Supplementary to figure 3A, B. Immunofluorescence staining for TF (green) and CitH3 (red) in **(Α)** neutrophils isolated from OA and RA SF, as well as **(Β)** peripheral blood neutrophils from OA, and RA patients. Panel (B) includes an isotype control to assess non-specific background staining. One representative example from three independent experiments performed with different donors is shown. Confocal microscopy; magnification: 60x; scale bar: 10 μm. TF, tissue factor; CitH3, citrullinated histone H3; OA, osteoarthritis; RA, rheumatoid arthritis; SF, synovial fluid.

**Supplementary Table**

|  | **Ctrls** | **OA** | **RA** |
| --- | --- | --- | --- |
| Number of samples | 5 | 8 | 9 |
| Per cent female | 80% | 87.5% | 87.50% |
| Age mean | 27 | 66.7 | 63 |
| Age range | (21; 35) | (60; 73) | (50; 78) |
| Per cent RF positive | nd | nd | 3 |
| Per cent ACPA positive | nd | nd | 9 |
| CRP mean (mg/dl) | 0.64 | 1.73 | 4.85 |
| ESR mean (mm/hr) | 19.2 | 22.2 | 40.8 |
| Patients on corticosteroids | nd | nd | 4 |
| Mean steroid dose (prednisolone equivalent) | nd | nd | 5mg/d |
| Patients on DMARDs | nd | nd | 5 |
| Methotrexate | nd | nd | 3 |
| Leflunomide | nd | nd | 3 |
| Hydroxychloroquine | nd | nd | 1 |
| Patients on biologicals | nd | nd | 1 |
| Tumour necrosis factor inhibitors | nd | nd | 2 |
| DAS28 | nd | nd | 4.9 |
| SF samples | nd | 8 | 9 |
| Isolation of FLS | 5 | 5 | 5 |

All numbers represent absolute values, except in the gender row, where they indicate the percentage of female patients. DMARDs, disease-modifying antirheumatic drugs; Ctrls, control subjects; nd, not defined; OA, Osteoarthritis; RA, Rheumatoid arthritis; DAS28, Disease Activity Score based on 28 joints; higher scores indicate greater rheumatoid arthritis activity; FLS, Fibroblasts-like synoviocytes; SF, synovial fluid; CRP, c-reactive protein; ESR, erythrocyte sedimentation rate.
